# How well do face masks protect the wearer compared to public perceptions?

**DOI:** 10.1101/2021.01.27.21250645

**Authors:** Eugenia O’Kelly, Anmol Arora, James Ward, P John Clarkson

**Author notes:** **Corresponding Author**: Eugenia O’Kelly, *Mailing Address*: USA: 999 Green St, Apt 1505, San Francisco, CA USA 94133, *E-mail. **Author details**: Eugenia O’Kelly, BA (Hon):, Anmol Arora, BA (Hons):, James Ward, PhD:, P John Clarkson, ScD. Funding*: This research received no specific grant from any funding agency in the public, commercial or not-for-profit sectors.

## Abstract

**Introduction:** There is a growing body of evidence to support the wearing of face masks to reduce spread of infectious respiratory pathogens, including SARS-CoV-2. However, the literature exploring the effectiveness of homemade fabric face masks is still in its infancy. Developing an evidence base is an important step to ensure that public policy is evidence based and truly effective.

**Methods:** Two methodologies were used in this study: quantitative fit testing of various face masks to indicate their effectiveness and a survey of 710 US residents about their perceptions of face mask effectiveness. N95, surgical and two fabric face masks were tested on an individual twenty five times each using a TSI 8038+ machine. Our survey was distributed by Qualtrics XM, asking participants to estimate the effectiveness of N95, surgical and fabric face masks.

**Results and Discussion:** Our results indicate that fabric face masks blocked between 62.6% and 87.1% of fine particles, whereas surgical masks protected against an average of 78.2% of fine particles. N95 masks blocked 99.6% of fine particles. Survey respondents tended to underestimate the effectiveness of masks, especially fabric masks. Together these results suggest that fabric masks may be a useful tool in the battle against the COVID-19 pandemic and that increasing public awareness of the effectiveness of fabric masks may help in this endeavour.

## Introduction

Since the beginning of the COVID-19 pandemic, a key concern from members of the public has been to what degree their face masks provide protection against small particles. Whilst the wearing of face masks has become either mandated or advised worldwide, there remains a notable shortage of studies assessing the effectiveness of masks, particularly those which are not traditionally used in clinical practice. N95, FFP3, and other hospital-grade masks are tested on each individual wearer to ensure that they can block a high percentage of particles which may cause the wearer harm. Non-medical grade masks, including surgical and homemade fabric masks, have no such ratings to enable wearers to estimate the filtration ability of their masks.

While early research has been conducted on both the ability of face mask fabrics to filter particles,^1^ and on the fit factor of non-medical grade masks,^2^ data illustrating the protection of non-medical grade masks is still lacking. The degree of public trust in the wearing of face masks remains unclear. It is also unclear to what extent perceived mask performance of different mask types drives mask selection amongst the public. Developing a body of literature assessing these face masks is an important step to ensure that public policy is science-based and that suitable policies are being propagated.

## Methods

To ascertain how successful a range of non-medical grade masks are at protecting the wearer, quantitative fit testing was performed with a TSI 8038+ machine. Quantitative fit testing is a reliable method of assessing the effectiveness of masks and is routinely used in practice to assess the fit of N95 and FFP3 masks before clinical use. Rather than calculating a fit factor using the US Occupational Health and Safety Administration (OHSA) formula, the machine was set to take air samples from both outside and inside the mask in short fifteen-second intervals. The number of particles, with a size range from 0.02 microns to over 1 micron, in these air samples were compared to derive the filtration efficiency. For reference, the size of the SARS-CoV-2 viral particle is roughly 0.12 microns.^3^ Twenty-five tests were performed on each mask on one volunteer.

In addition to these experiments, a survey was distributed by Qualtrics XM (December 2020 - January 2021) to a sample aimed to be a representative of the US general population. To ascertain the public’s perception of mask effectiveness at filtering harmful particles, 710 US residents were asked to guess approximately what percentage of particles N95 masks, surgical masks, and fabric masks protect the wearer from. Participants were given two example pictures of each mask type and were asked to assign N95, surgical, and fabric masks to one of five categories of filtration efficiency: 0-25% of particles, 25-50% of particles, 50-75% of particles, 75-95% of particles, and over 95% of particles.

## Results

Results showed that non-medical grade masks were effective in blocking over 50% of airborne particles, with the simple fabric mask and surgical mask blocking between 50% and 75% of particles (Fig.1). Fabric mask filtration was seen to differ significantly based on the design of the mask. A simple two-layer thin cotton fabric mask blocked an average of 62.6% of particles while the more sophisticated fabric mask with panels to cover the nose and chin and a PM2.5 filter blocked an average of 87.1% of particles. This indicates the construction of fabric masks is key to their effectiveness. The N95 mask blocked approximately 99.6% of particles.

**Figure 1:**
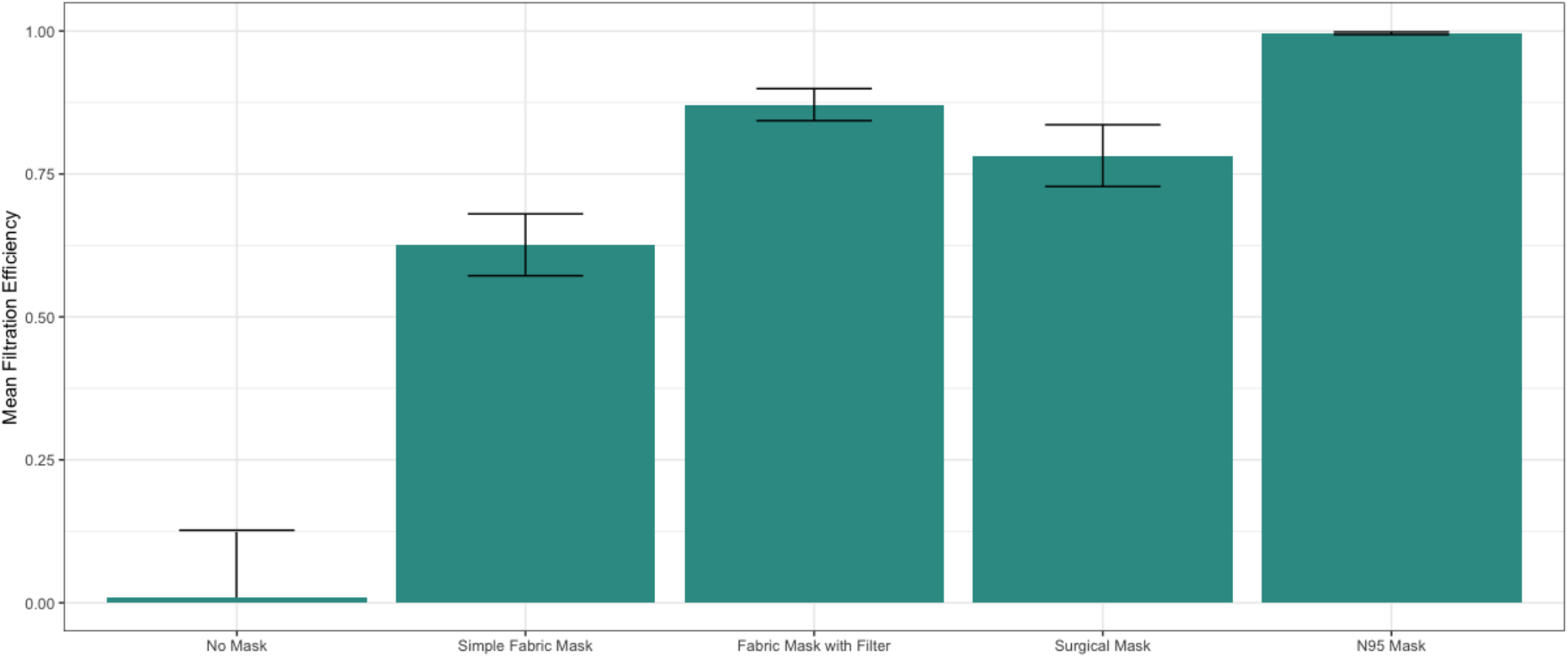
Comparison the mean filtration efficiencies measured for four mask types: a simple two-layer fabric mask, a more sophisticated mask with panels to cover the nose and chin and a PM2.5 filter, a standard surgical mask and an N95 mask. Error bars indicate standard deviation above and below the mean. To accurately compare the state of not wearing a mask, a special headpiece was designed to hold the mask sampling tube the same distance from the mouth as it would be when a mask was worn.

**Figure 2:**
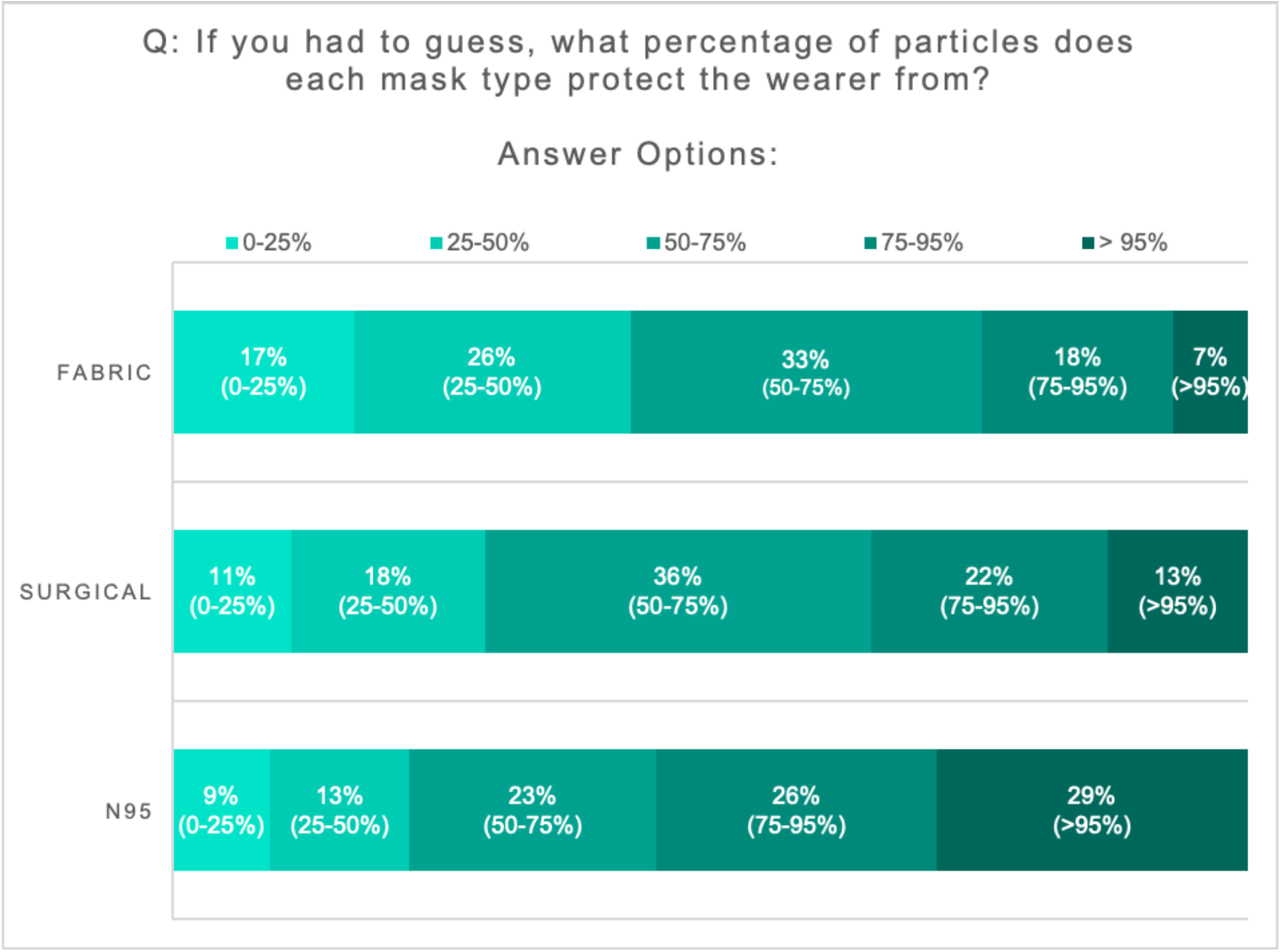
Results of 710 US residents estimating approximately what percentage of particles different masks protect the wearers from.

A non-medical grade surgical mask, worn tight to the face by adjusting the straps, filtered an average of 78.2% of particles. While none of these masks compared to the 99.6% measured filtration efficiency of the N95 mask tests, non-medical grade masks still managed to provide high levels of protection (62.6-87.1%) compared to not wearing a mask (1.0%).

The survey results found that 78% of respondents believed N95 masks protected from over 50% of particles, but only 29% believed the masks protected from over 95% of particles. When considering surgical masks, 29% of respondents believed the mask filtered less than 50% of particles, with the greatest proportion, 36%, believe the mask filtered between 50% and 75% of particles. Participants judged the fabric masks the least effective, with 43% believing they filtered less than 50% of particles. Surgical masks and fabric masks had similar proportions of respondents guessing a filtration level of 50-75%, with 36% in the case of surgical masks and 33% in the case of fabric masks.

## Discussion

Overall, our results suggest that even simple fabric masks may be able to protect the wearer from fine particles the size of the SARS-CoV-2 viral particle. In doing so, these findings add to the growing body of evidence suggesting that mask wearing may play an important role in the global response to the COVID-19 pandemic. Beyond simply reducing transmission, there is also valid reason to believe that protecting civilians from such percentages of pathogens may lead to improved clinical outcomes. It has been hypothesised that the severity of COVID-19 illness may be related to the size of the viral inoculum a subject is exposed to, and that mask-wearing may increase the likelihood of a milder disease course.^4,5^

Overall, the public perception of mask effectiveness was reasonably accurate in the case of fabric masks and surgical masks. Participants were more likely to underestimate the filtration of masks than overestimate the filtration. Relatively few participants overestimated the effectiveness of face masks, especially for fabric masks. On the other hand, underestimation was common. Over 20% of participants considered N95 masks to be less than 50% effective, when in actuality these masks were found to be over 99% effective, masks which as the name suggests, are certified to be over 95% effective if they fit. Similarly over 40% of participants thought that fabric masks were less than 50% effective compared with the 62.6% measurement for the simple fabric mask we tested.

There was a clear separation in the results of perceived effectiveness between masks which are used in clinical practice and fabric masks. In order to sustain supply of face masks for hospitals and front-line workers, it is important to emphasise that fabric masks are indeed effective and in some cases can outperform surgical masks, especially when more complex designs are used. Fabric masks also carry advantages of being reusable, often being less expensive and having lower environmental impacts than clinically used masks. Overall, the results of this study suggest that fabric face masks may be a useful tool in the COVID-19 pandemic arsenal, and information campaigns may be advisable to increase awareness of the benefits that they offer.

## Data Availability

All relevant data are contained within the article.

## References

1. O’Kelly E, Pirog S, Ward J, Clarkson PJ. Ability of fabric face mask materials to filter ultrafine particles at coughing velocity. BMJ Open. 2020 Sep 1;10(9):e039424.

2. O’Kelly E, Arora A, Pirog S, Ward J, Clarkson PJ. Comparing the Fit of N95, KN95, Surgical, and Cloth Face Masks and Assessing the Accuracy of Fit Checking. medRxiv. 2020. doi.org/10.1101/2020.08.17.20176735

3. Bar-On YM, Flamholz A, Phillips R, Milo R. SARS-CoV-2 (COVID-19) by the numbers. Eisen MB, editor. eLife. 2020 Mar 31;9:e57309.

4. Gandhi M, Beyrer C, Goosby E. Masks Do More Than Protect Others During COVID-19: Reducing the Inoculum of SARS-CoV-2 to Protect the Wearer. J GEN INTERN MED. 2020 Oct 1;35(10):3063–6.

5. Guallar MP, Meiriño R, Donat-Vargas C, Corral O, Jouvé N, Soriano V. Inoculum at the time of SARS-CoV-2 exposure and risk of disease severity. International Journal of Infectious Diseases. 2020 Aug 1;97:290–2.

